# Mapping the trajectory of psychotic symptoms and their interaction with antipsychotic treatment: a longitudinal network intervention study

**DOI:** 10.1101/2025.05.28.25328478

**Authors:** Pierfrancesco Sarti, Giacomo Cecere, Victoria Edkins, Wolfgang Omlor, Johanna M.C. Blom, Philipp Homan

## Abstract

Antipsychotic efficacy in schizophrenia spectrum disorders (SSD) is commonly evaluated using static measures that fail to capture the dynamic evolution of symptoms and the unfolding impact of treatment over time. Network Intervention Analysis (NIA) is a novel approach that models pharmacological treatments as active nodes within longitudinal symptom networks, capturing both direct and indirect treatment effects. This study aimed to investigate how the receptor-binding profile of antipsychotics influence symptom trajectories over six weeks. NIA was used to characterise the evolving impact of treatment with muscarinic antagonists, serotonergic/dopaminergic antagonists, and adrenergic agents with low dopaminergic antagonism within dynamic symptom networks. We hypothesised that NIA would reveal distinct patterns of symptom change, reflecting the pharmacodynamic mechanisms specific to each drug class. Forty-seven patients with SSD underwent baseline assessments including neuropsychological tests and five symptom rating scales from the Manual for the Assessment and Documentation of Psychopathology in Psychiatry (AMDP). They were then followed weekly for six additional weeks evaluating them with the AMDP-based symptom ratings, providing a comprehensive, standardised, and fine-grained measure of psychopathological changes. Muscarinic antagonists initially targeted self-disorder, then shifted to delusions, reducing symptom interconnectivity and increasing network resilience. Serotonergic/dopaminergic antagonists primarily influenced hallucinations but showed a late stage rebound, with increased network density and reduced treatment influence. Adrenergic agents exhibited a stabilising effect, preserving network structure with minimal symptom reduction. These findings demonstrate the utility of NIA in capturing the temporal dynamics of antipsychotic effects based on receptor affinity, supporting the development of phase-specific, network-informed, and personalised interventions.

## 1. Introduction

Antipsychotics are the mainstay of treatmen–t in schizophrenia spectrum disorders (SSD), playing a crucial role in managing symptoms and preventing further clinical deterioration (Sampogna et al., 2023). These medications are typically categorised as belonging to one of three classes: First-generation antipsychotics (FGAs) primarily block dopamine receptors; second-generation antipsychotics (SGAs) act on both dopamine and serotonin receptors; and third generation antipsychotics are partial dopamine agonists, which modulate dopamine activity locally (Kaar et al., 2019; Kapur et al., 2004).

This classification system is of limited utility in the selection of an appropriate antipsychotic medication and has drawn criticism for failing to map either the medications’ pharmacological or clinical effects (McCutcheon et al., 2024). The choice of medication involves a complex clinical decision-making process shared by clinician and patient and influenced by interrelated factors including clinician expertise, patient characteristics and medication properties (Fiorillo et al., 2020). The clinician must review the patient’s symptoms, consider patient-specific attributes (such as age, illness duration, comorbidities, and previous medication experiences), and evaluate medication characteristics (for example side effect profiles, efficacy potential, and the possibility of switching to a long acting injectable). Each of these factors significantly impact treatment outcomes and relapse risk (Aprile et al., 2025; Kane et al., 2020, 2003; Siafis et al., 2022).

Although FGAs and SGAs have been extensively studied (Keefe et al., 2007), their precise effects over time and across symptom domains (positive, negative, cognitive) remain unclear, with research yielding complex and sometimes conflicting results (Lee et al., 2024; Woodward et al., 2007). For instance, a meta-analysis by Cheuk et al., (2024) found that the SGA Clozapine improved cognitive symptoms, while a more recent meta-analysis by Feber et al. (2025) reported that Clozapine, like FGAs, has a detrimental effect on cognition. Further evidence of the complexity of antipsychotics’ effects comes from two major network meta-analyses assessing the efficacy and side effects of FGAs and SGAs (Huhn et al., 2019; Leucht et al., 2023). These studies highlight that a focus on efficacy alone is not sufficient when selecting an antipsychotic; the side effect profile and its variability is also relevant to medication selection (Neumeier et al., 2021). Findings such as these exemplify why antipsychotic selection remains challenging despite decades of research and underscore the importance of individualised treatment approaches. There are numerous internal and external variables that potentially contribute to antipsychotics’ effects. Clinicians must integrate these patient-specific factors with evolving evidence to achieve optimal patient outcomes (Keefe et al., 2007).

To better capture the heterogeneity of both biochemical and symptomatologic effects of antipsychotics, an additional classification approach has been proposed: McCutcheon et al. (2023) introduced a data-driven taxonomic approach to classify antipsychotics. The classification was based on effects and side effects according to antipsychotics’ receptor affinity (muscarinic, adrenergic/low dopaminergic, serotonergic/dopaminergic and dopaminergic). This receptor-based classification predicted clinical effects better than existing classification schemes, with a statistically significant prediction of out-of-sample clinical effect profiles, superior to the atypical/typical classification (McCutcheon et al., 2023).

Despite this effort to refine antipsychotic classification, current pharmacological research still struggles to fully capture the complexity of SSD treatment (Huhn et al., 2019; Winkelbeiner et al., 2019; Zhang et al., 2021). While receptor-based classifications offer better predictions of clinical effects, research methodologies continue to rely on static assessments and group-level comparisons, often overlooking the dynamic and individualised nature of treatment response. Traditional study designs, such as randomised controlled trials (RCTs), assess efficacy at the population level, assuming relatively stable treatment effects across patients. However, schizophrenia presents substantial heterogeneity not only in neurobiology but also in symptom expression and progression, making it crucial to move beyond these methodologies. A more dynamic, personalised approach—accounting for evolving symptom interactions and individual treatment trajectories—could enhance understanding of antipsychotic effects and ultimately improve treatment strategies.

Network analysis (NA) has emerged as a powerful methodological framework for studying psychiatric disorders. NA conceptualises these conditions as dynamic systems in which symptoms interact and reinforce each other (Borsboom, 2017). This allows for a more nuanced understanding of psychopathology, emphasising the importance of symptom connectivity and centrality rather than relying solely on categorical diagnoses or sum scores. By mapping symptoms and their interrelations, NA enables researchers to identify key nodes within the network that may act as pivotal drivers of disorder progression (Borsboom and Cramer, 2013).

Within this broader framework, network intervention analysis (NIA) is an advanced analytical tool designed to study the effects of pharmacological and non-pharmacological treatments on symptom networks over time. NIA captures the temporal dynamics of symptom change, allowing differentiation between direct and indirect treatment effects (Blanken et al., 2019).

In SSD, the complexity of clinical and neurobiological phenotypes, along with the relationship between individual characteristics, clinical manifestation and pharmacological treatment, pose significant challenges to understanding disease mechanisms and therefore the ability to optimise treatment strategies. NIA offers a promising approach to simultaneously address these complex interactions (Monteleone et al., 2021). Unlike traditional approaches —such as unidimensional models, fixed-time assessments, and categorical treatment outcomes (e.g., response vs. non-response)— which often rely on overall severity scores and assume linear treatment effects, NIA can track how specific symptoms shift in response to treatment across multiple time points. With this information, researchers can determine which symptoms serve as key drivers of network reorganisation. This enables the identification of treatment-sensitive nodes—symptoms that, when effectively managed, will lead to widespread improvements across the network. Additionally, by leveraging centrality statistics—such as strength, betweenness, and expected influence—NIA helps to pinpoint symptoms that are most influential in sustaining psychopathological states, guiding the prioritisation of intervention strategies (Boschloo et al., 2019).

NIA has been applied successfully to major depression to examine differences in how cognitive and behavioural therapy affect insomnia (Lancee et al., 2022), and to identify the distinct symptom targets of antidepressants and antipsychotics (Guerrera et al., 2023). Additionally, it has been used to explore anxiety-related issues among cancer survivors (Fishbein et al., 2023). Most published studies have focused on mapping the interactions between psychotherapy treatments and the temporal dynamics of symptoms. Relatively few have applied it to pharmacological treatment (Guerrera et al., 2023). A recent study applied NIA to examine the effects of the experimental drug Ruloperidon (James et al., 2024). The authors highlighted NIA’s potential in capturing both direct and indirect treatment effects on negative symptoms over time. Specifically, NIA indicated that a particular dose of Roluperidone had a direct effect on avolition, which in turn influenced the broader network of negative symptoms. This suggests that changes in patients’ negative symptomatology were mediated by the drug’s direct effects on avolition rather than other symptoms.

The potential role and application of NIA in SSD remain largely unexplored, but there have been relevant attempts to explore the longitudinal impact of pharmacological treatment on symptoms using networks. For instance, Zamani et al. (2017) examined network changes in the Clinical Antipsychotic Trials of Intervention Effectiveness (CATIE) (Stroup et al., 2003; Zamani, 2017) sample over an 18-month period. They found increased network connectivity in treatment-responsive patients, while treatment-resistant individuals showed no significant change. Although the sample was large and clinically relevant, the study’s two-time-point design offers a relatively static perspective, overlooking the non-linear and evolving nature of treatment effects across time. Sun et al. (2023) investigated network changes in a large sample of acutely psychotic patients over four bi-weekly time points. They found that antipsychotic treatment significantly altered the structure of symptom networks in schizophrenia, specifically by reducing overall network strength and the number of connections between nodes. The approach used provided valuable insight into the temporal modulation of symptom interrelations. However, it treated pharmacological interventions as stratification factors rather than active components of the network, limiting the capacity to model direct versus indirect treatment effects.

In this study, we build on these previous efforts by applying NIA to model treatment as an active node within the network, allowing us to examine the evolving interplay between symptoms and pharmacological interventions in patients with SSD. We hypothesised that NIA would reveal specific patterns of symptom change for each antipsychotic class depending on their unique pharmacodynamic mechanisms of action. By tracking symptom-treatment relationships across seven weekly time points, we aimed to capture both cross-sectional differences in patients’ responses to drugs within each class and longitudinal variations in these responses.

This approach, grounded in precision medicine, sought to identify key network nodes that drive treatment response, offering actionable and nuanced understanding of how different receptor profiles influence symptom trajectories. In doing so, it holds the potential to inform targeted pharmacological and non-pharmacological interventions, which optimise treatment efficacy while minimising adverse effects.

## 2. Experimental procedures

### 2.1 Participants

The patients included in this study were part of a larger case-control study (Striatal Connectivity in Psychosis - STRICON) conducted at the University Hospital of Psychiatry in Zurich and approved by the Swiss Ethics Commission (ID 2020-00277). We analysed the data collected from 47 patients recruited for the STRICON study. The inclusion and exclusion criteria of this study were the same those for the larger STRICON study which were as follows:

**Inclusion Criteria**

- Diagnosis of a psychotic disorder according to the International Classification of Diseases (ICD-10)
- In remission following a psychotic episode, defined as having current positive symptoms rated as 3 (mild) or lower on the Brief Psychiatric Rating Scale
- Undergoing both psychopharmacological treatment and psychotherapeutic care
- Aged between 18 and 40 years
- Able to understand the study details and provide informed consent

**Exclusion criteria**

- Serious medical, organic, or neurological conditions that could explain the psychiatric symptoms
- Use of medications with antipsychotic properties for non-psychiatric reasons
- Ongoing drug dependency or substance abuse
- Significant risk of suicidal behaviour
- Undergoing treatment with clozapine
- Current use of benzodiazepines or sedating medications
- Pregnancy

### 2.2 Study design and procedure

This was a naturalistic, longitudinal observational study. All assessments and testing were conducted in a controlled laboratory setting within the University Hospital of Psychiatry in Zurich by trained clinicians. Patients had already been receiving pharmacological treatment for two weeks and their assignment to one of three antipsychotic treatment groups was based on clinical evaluation and conducted through a shared decision-making process. The three treatment groups were:

- Group 1 (muscarinic M2–M5 antagonists): olanzapine, quetiapine
- Group 2 (serotonergic and dopaminergic antagonists): paliperidone, risperidone
- Group 3 (adrenergic agents with low dopaminergic antagonism): aripiprazole, lurasidone, brexpiprazole, cariprazine

After providing informed consent, participants underwent a comprehensive psychopathological assessment at baseline (T0), conducted in close temporal proximity to neuroimaging (MRI) sessions (the neuroimaging data were not analysed in this study). Of the psychological tests conducted, the following were used in this study:

- The Positive subscale of the Positive and Negative Syndrome Scale (PANSS P) was used to measure positive symptomatology (e.g., hallucinations, delusions). This subscale consists of 7 items, each rated on a Likert scale from 1 to 7, where higher scores indicate greater symptom severity (Kay et al., 1987).
- The Brief Negative Symptom Scale (BNSS) assesses negative symptoms in schizophrenia. It measures six domains: blunted affect, alogia, anhedonia, asociality, distress, and avolition. The items in each domain are coded using a 7-point Likert scale ranging from absent (0) to severe (6) (Kirkpatrick et al., 2011).
- The Brief Psychiatric Rating Scale (BPRS) is used to assess the severity of psychotic and affective symptoms. The scale comprises 18 items, each rated on a Likert scale from 1 to 7 scale, where higher scores indicate greater symptom severity (Overall and Gorham, 1962).
- The Snaith-Hamilton Pleasure Scale (SHAPS) was administered to specifically measure anhedonia. The scale consists of 14 items; each rated on a dichotomous scale (Snaith et al., 1995).
- The MATRICS Consensus Cognitive Battery (MCCB) is a standardised test battery for assessing cognition in schizophrenia (Nuechterlein et al., 2008). Six of the battery’s seven domains were used: processing speed, attention/vigilance, working memory, verbal learning, visual learning, reasoning/problem solving. Raw scores are reported as T-scores (mean = 50, SD = 10), with a composite score reflecting overall performance. T-scores (percentile equivalents) were transformed into equivalent scores from 0 to 4 according to the following breakdown: 0^th^ - 4^th^ = 0; 5^th^ - 15^th^ = 1; 16^th^ - 84^th^ = 2; 85^th^ - 94^th^ = 3; 96^th^ - 100^th^ = 4.
- The Manual for the Assessment and Documentation of Psychopathology in Psychiatry (AMDP) is a standardised and internationally used system for the description and assessment of psychopathological findings. It includes several rating forms to code the severity of each symptom on a scale from 0 (“absent”) to 3 (“severe”), allowing both qualitative descriptions and quantitative scoring of psychopathological states. The AMDP rating forms for formal thought disorder, fears and compulsions, derealisation, hallucinations, and self-disorder were used (Stieglitz et al., 2017).

Follow-up assessments carried out by trained hospital psychiatrists were conducted weekly for six consecutive weeks (T1 – T6). Symptom severity was rated using AMDP during structured clinical interviews.

### 2.4 Statistical Analysis

All statistical analysis, network models, and network visualisations for the three treatment classes at the seven time points were conducted using R software version 4.4.2. Bar charts were created using GraphPad Prism v. 9.00e.

To examine the pattern of missing assessment data, an exploratory analysis and a graphical visualisation of the missing value distribution were performed, followed by imputation of missing values using the predictive mean matching (PMM) method (Bailey et al., 2020). Imputation was executed with the *mice* package (Buuren and Groothuis-Oudshoorn, 2011). Five imputed datasets were generated, and the quality of the imputation was assessed by comparing the distribution of observed and imputed data. The highest quality dataset was selected to conduct the subsequent analyses.

To evaluate the distribution of variables in the completed dataset, the Shapiro-Wilk normality test was conducted for each variable. Subsequently, a correlation analysis was performed including the variables measured at T0. The correlation matrix was calculated using Spearman’s method.

At T0, differences in the means of the three patient groups for all tests and questionnaires administered during the neuropsychological assessment were explored using the Kruskal-Wallis test. If significant, Dunn’s test was performed as a post-hoc analysis.

To assess the changes from T0 to T6 in the mean scores for each of the five AMDP rating forms and to estimate the average trajectory of symptom severity, a linear mixed effects model for each pharmacological class was implemented. The outcome variable was the weekly average AMDP symptom score. Time (in weeks) was included as a fixed effect. Random intercepts and slopes for time were specified at the subject level to account for individual differences in baseline severity and change over time. Models were estimated using restricted maximum likelihood (REML) in the lme4 package in R.

For the NA, we employed a multi-group network analysis over seven time points (T0 to T6) using mixed graphical models (MGM) to construct networks, assess predictability, and compute centrality measures (Sedgewick et al., 2016). For each time point, a network was estimated using the *mgm* package (Haslbeck and Waldorp, 2020), applying the extended Bayesian information criterion (EBIC) for model selection to balance model fit and complexity (Chen and Chen, 2008). The AND rule for regularisation was used, ensuring that an edge between two nodes was included only if both directed edges were nonzero. Ridge regression was implemented as a form of L2 regularisation to prevent overfitting by shrinking coefficients towards zero. The input data comprised five continuous variables (the mean scores of AMDP rating forms) and one dichotomous variable (presence or absence of each antipsychotic class). Analyses were performed separately for each antipsychotic class, using the other two classes as comparators.

Network graphs were plotted using the *qgraph* package with a circular layout to standardise visualisation (Epskamp et al., 2012). Predictability was assessed to determine how well the value of a node could be predicted based on the rest of the network (Haslbeck and Waldorp, 2018). For continuous variables, the coefficient of determination (R^2^) was used as a measure of variance explained. For categorical variables, normalised classification accuracy (nCC) was used to evaluate the predictability of categorical states. Predictability was then extracted for graphical representation, with values overlaid on a pie chart in the network visualisation. Each network was converted into an igraph object to calculate the connection density.

Four centrality metrics were computed: strength, betweenness, closeness, and expected influence (EI). EI was chosen as the primary centrality metric for interpretation, as it retains the directionality of effects—unlike strength—and has been shown to be more reliable than betweenness in psychopathology networks Robinaugh et al. (2016), making it especially suitable for psychological and clinical applications. For visualisation purposes, EI values were normalised between 3 and 8 to prevent extreme variations in node sizes. In the tables they were kept within the range 0 to 1. Finally, to assess the robustness of network estimates, a bootstrap procedure was performed at each time point for each treatment condition using the *bootnet* package with 1000 nonparametric resamples (Epskamp et al., 2018). Stability of key network parameters, including strength, expected influence, betweenness, and edge weights, was evaluated. Bootstrapped centrality metrics were plotted to visualise their variability, and edge stability was examined using confidence intervals.

All R code used for the data imputation analyses and network analyses is provided as R-Markdown.

## 3. Results

The descriptive statistics for the sample and for the three groups separately at T0 are presented in Table 1.

**Table 1:** Demographic characteristics and neuropsychological assessment scores of the sample at Time 0, classified according to the receptor affinity of the medication received. Biological sex is reported as frequencies. Disease duration is reported as mean, minimum and maximum values. Other continuous and ordinal variables are reported as means and standard deviations. The H statistics and their relative p-values, resulting from Kruskal-Wallis tests used to compare the three patient groups’ neuropsychological test values are also reported. A p-value of “ns” indicates not significant, “*” indicates p < .05. Note: MATRICS = MATRICS Consensus Cognitive Battery; SHAPS = Snaith-Hamilton Pleasure Scale; BPRS = Brief Psychiatric Rating Scale; PANSS P = Positive symptoms subscale of the Positive and Negative Syndrome Scale; BNSS = Brief Negative Symptom Scale.

| Variable | Muscarinic | Serotonergic/Dopaminergic | Adrenergic/Lo<br>w<br>dopaminergic | Total sample | Kruskal-Wallis' H | p-value |
| --- | --- | --- | --- | --- | --- | --- |
| **Numerosity** | 19 | 18 | 10 | 47 | \ | \ |
| **Sex** | M = 14; F = 5 | M = 14; F = 4 | M = 7; F = 3 | M = 35; F = 12 | \ | \ |
| **Age** | 24.68 (5.37) | 27.33 (6.07) | 28.80 (7.15) | 26.92 (6.39) | 3.16 | ns |
| **Years of education** | 12.37 (1.45) | 11.61 (1.75) | 12.50 (1.35) | 12.11 (1.45) | 2.73 | ns |
| **Disease duration in months; min-** | 11.79; 1-33 | 19.83; 1-86 | 38.4; 2-192 | 19.21; 1-192 | 3.38 | ns |
| max |  |  |  |  |  |  |
| **Neuropsychological Assessment at T0** |  |  |  |  |  |  |
| **MATRICES** |  |  |  |  |  |  |
| - Attention/vigilance | 0.89 (0.94) | 0.72 (0.89) | 1.4 (0.07) | 1.02 (1.04) | 2.948 | ns |
| - Processing speed | 1.21 (0.79) | 0.66 (0.91) | 1.2 (1.03) | 1.08 (0.99) | 4.593 | ns |
| - Working memory | 1.47 (0.84) | 1.16 (0.86) | 1.5 (0.85) | 1.4 (0.87) | 1.642 | ns |
| - Verbal learning | 1.84 (0.83) | 1.27 (1.02) | 2.3 (1.16) | 1.71 (1.04) | 6.426 | * |
| - Problem solving | 1.42 (0.90) | 1.27 (0.96) | 1.6 (0.97) | 1.46 (0.98) | 0.801 | ns |
| - Visual learning | 1.57 (0.77) | 1.16 (0.86) | 1.5 (0.85) | 1.5 (0.85) | 3.002 | ns |
| **SHAPS** | 5.63 (2.19) | 6.39 (2.28) | 5.8 (3.79) | 5.58 (2.77) | 1.522 | ns |
| **BPRS** | 41.58 (8.73) | 37.5 (5.88) | 40.3 (16.91) | 39.1 (1.04) | 4.043 | ns |
| **PANSS P** |  |  |  |  |  |  |
| - Total score | 13.52 (5.12) | 13.72 (4.79) | 18.33 (6.88) | 14.46 (5.53) | 3.288 | ns |
| - Delusions | 2.79 (1.4) | 2.94 (1.47) | 3.8 (1.69) | 3.06 (1.55) | 2.606 | ns |
| - Concept. Disorg. | 2.11 (1.29) | 2.22 (1.48) | 2.4 (1.35) | 2.12 (1.32) | 0.447 | ns |
| - Hallucin. behav. | 2.37 (1.34) | 2.61 (1.5) | 3.7 (1.34) | 2.62 (1.46) | 5.233 | ns |
| - Excitement | 1.47 (0.7) | 1.11 (0.32) | 1.7 (1.06) | 1.37 (0.69) | 4.387 | ns |
| - Grandiosity | 1.37 (0.68) | 1.56 (0.92) | 1.7 (1.25) | 1.56 (1.07) | 0.572 | ns |
| - Suspiciousness | 2.37 (1.21) | 2.28 (1.49) | 2.5 (1.58) | 2.35 (1.36) | 0.379 | ns |
| - Hostility | 1.05 (0.23) | 1 (0) | 1.2 (0.63) | 1.06 (0.31) | 1.671 | ns |
| **BNSS** |  |  |  |  |  |  |
| - Total score | 44.84 (13.01) | 41.66 (14.33) | 37.2 (19.41) | 42 (14.97) | 1.551 | ns |
| - Asociality | 6.42 (2.63) | 6.78 (2.6) | 5.5 (2.92) | 6.08 (2.75) | 2.272 | ns |
| - Avolition | 7.35 (2.55) | 6.28 (2.3) | 6.7 (3.59) | 6.6 (2.75) | 2.225 | ns |
| - Anhedonia | 12.42 (4.27) | 11.11 (4.16) | 9.3 (5.68) | 10.88 (4.67) | 2.823 | ns |
| - Distress | 3.79 (1.47) | 3.5 (1.42) | 2.5 (1.65) | 3.31 (1.52) | 5.043 | ns |
| - Blunted affect | 10.32 (4.42) | 9.72 (4.53) | 8.4 (5.23) | 9.4 (4.53) | 1.524 | ns |
| - Alogia | 4.37 (2.22) | 4.5 (2.09) | 4.8 (3.97) | 4.37 (2.51) | 0.298 | ns |

The three patient groups were comparable in terms of age. The ratio of males to females was 3:1 in the muscarinic and serotoninergic dopaminergic groups and 2:1 in the adrenergic low dopaminergic group.

Of the Kruskal-Wallis tests comparing mean neuropsychological assessment scores at T0, one was significant: A significant difference was found between the patient groups for the MATRICS verbal learning subtest score (H = 6.426, p = 0.042). Dunn’s post-hoc tests showed a significantly higher score in the adrenergic/low dopaminergic group compared to the serotoninergic/dopaminergic group (mean difference = 1.03 p = 0.049).

Participants’ positive symptomatology measured with the PANSS, ranged between mild and moderate on items related to delusions (mean = 3.06), conceptual disorganisation (mean = 2.12), hallucinatory behaviour (mean = 2.62), and suspiciousness (mean = 2.35) in all three groups. For negative symptomatology, the BNSS showed the highest scores in the anhedonia (mean = 10.88, range 0-18) and blunted affect (mean = 9.4, range 0-18) domains, indicating moderate to high levels of reduced pleasure and moderate impairment in emotional expression.

All three groups had MATRICS subscale scores that fell below the 46th percentile, meaning that they performed worse than 54% of the normative population. The greatest impairment was observed in attention and vigilance (mean equivalent score = 1.02), corresponding to the 25^th^ percentile, and in processing speed (mean equivalent score = 1.08), corresponding to the 27^th^ percentile.

Symptom trajectories over six weeks decreased across all pharmacological groups, though the magnitude and consistency of improvement varied. The muscarinic group (Figure 1A) showed the largest average weekly reduction in symptom severity (β = −0.030, SE = 0.0055), with a strong negative correlation between baseline severity and rate of improvement (r = −0.98). The serotonergic/dopaminergic group (Figure 2A) exhibited a more modest improvement (β = −0.016, SE = 0.0051), with less consistent symptom reductions and a weaker baseline-slope correlation (r = −0.25). The adrenergic/low-dopaminergic group (Figure 3A) had a similar average slope (β = −0.016, SE = 0.0073), and a strong baseline-slope correlation (r = −0.76). Interindividual variability in symptom trajectories was comparable across groups. Model summaries for overall trajectories and individual symptoms are reported in Tables 1 and 2 in the Supplementary Material.

**Figure 1:**
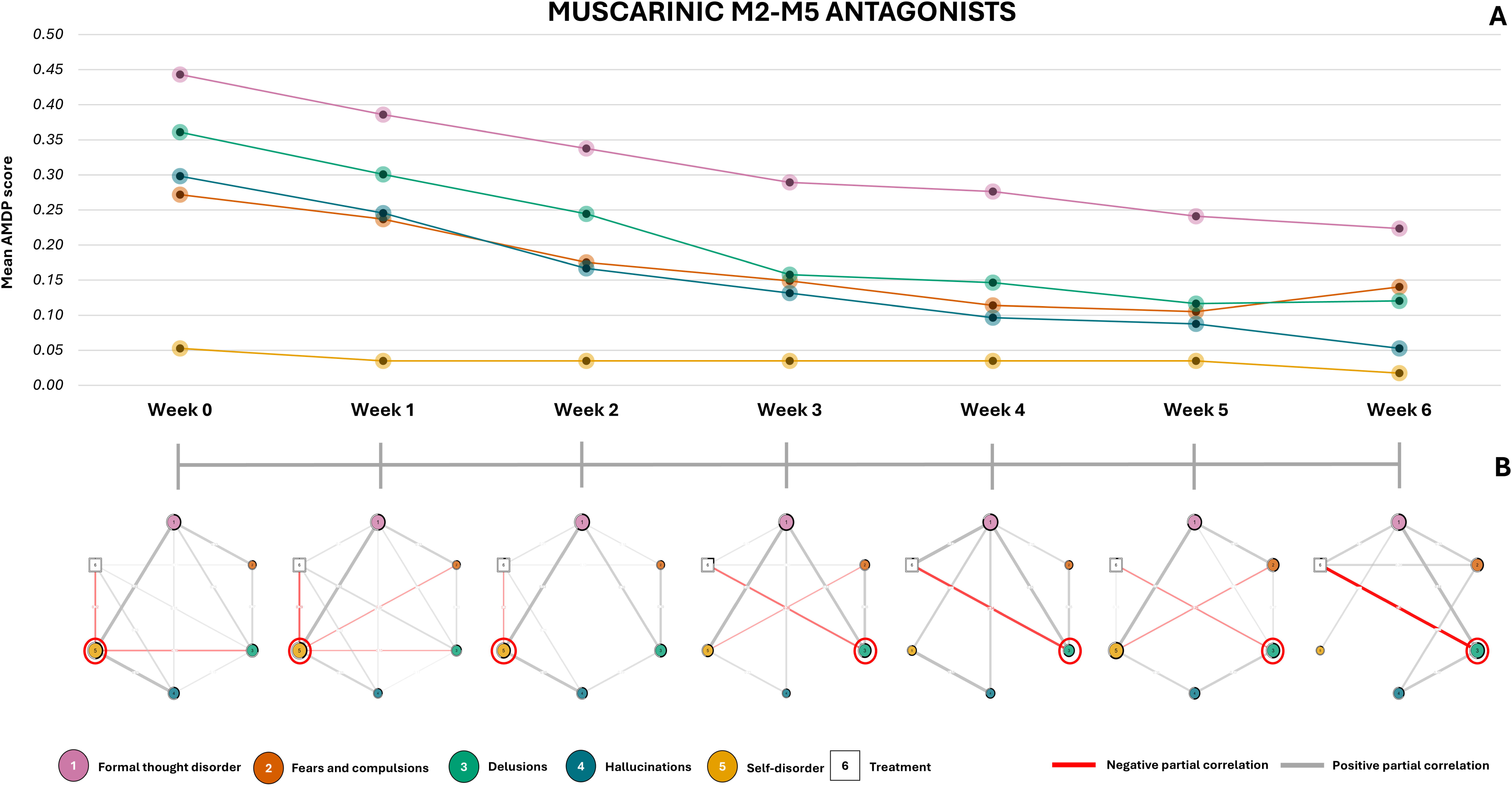
Part A, line graph showing the development of symptoms over time for each symptom category from the Manual for the Assessment and Documentation of Psychopathology in Psychiatry (AMDP) from T0 to T6 (given as mean of the AMDP rating forms). Part B, network models corresponding to each time point. Grey connections indicate positive partial correlations, while red connections indicate negative partial correlations. The ring around each node indicates its predictability value. The darker the ring, the higher the value. The size of the nodes is proportional to their expected influence value. The colour of the nodes corresponds to that of the lines in the Part A graph. The white square node represents the pharmacological treatment. The red circles mark the symptom nodes upon which the pharmacological treatment exerts a direct effect, contributing to their reduction.

**Figure 2:**
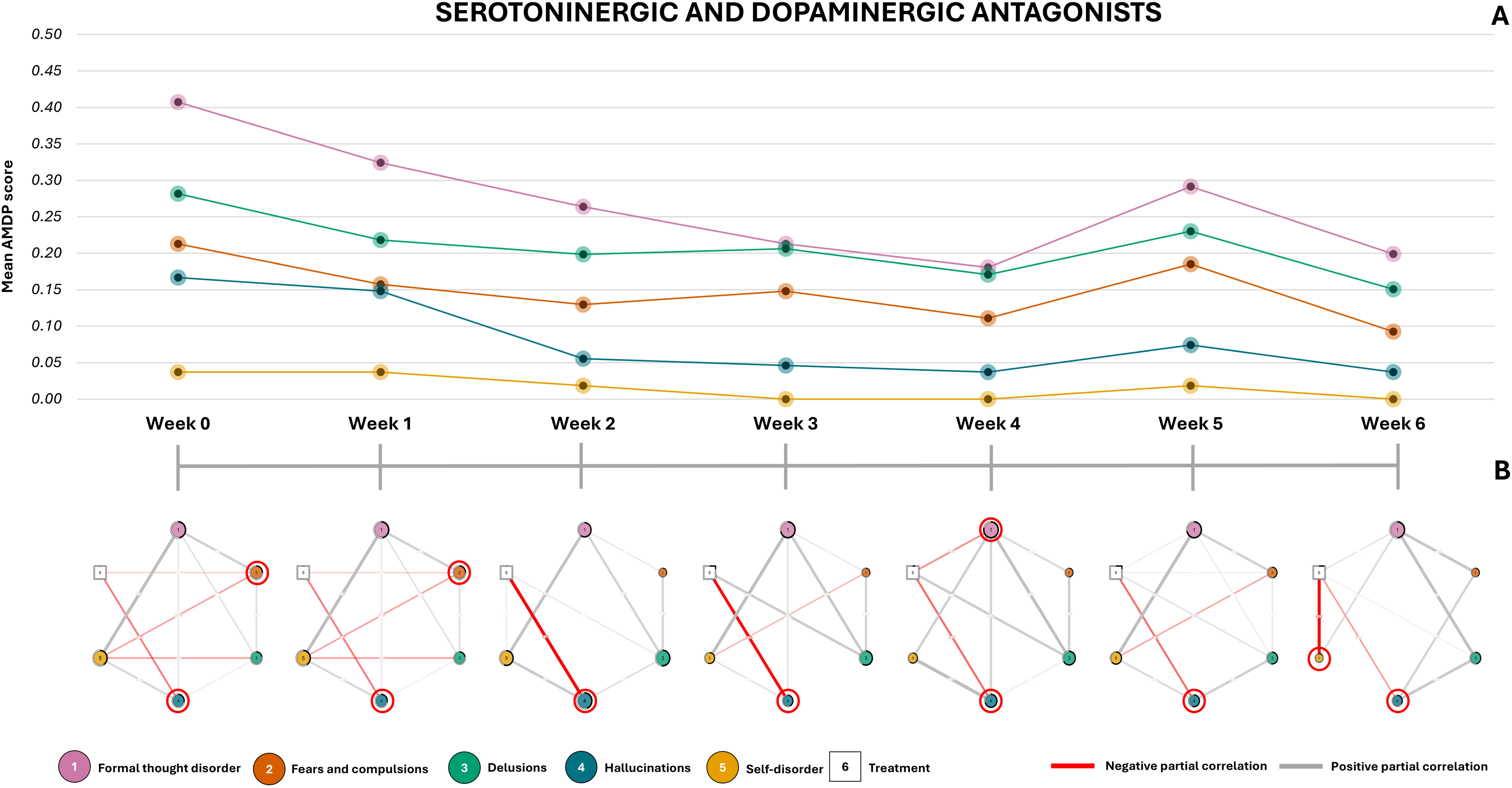
Part A, line graph showing the development of symptoms over time for the Manual for the Assessment and Documentation of Psychopathology in Psychiatry (AMDP) from T0 to T6 (given as mean of the AMDP rating forms). Grey connections indicate positive partial correlations, while red connections indicate negative partial correlations. The ring around each node indicates its predictability. The darker the ring, the higher the value. The size of the nodes is proportional to their expected influence value. The colour of the nodes corresponds to that of the lines in the Part A graph. The white square node represents the pharmacological treatment. The red circles mark the symptom nodes upon which the pharmacological treatment exerts a direct effect, contributing to their reduction.

**Figure 3:**
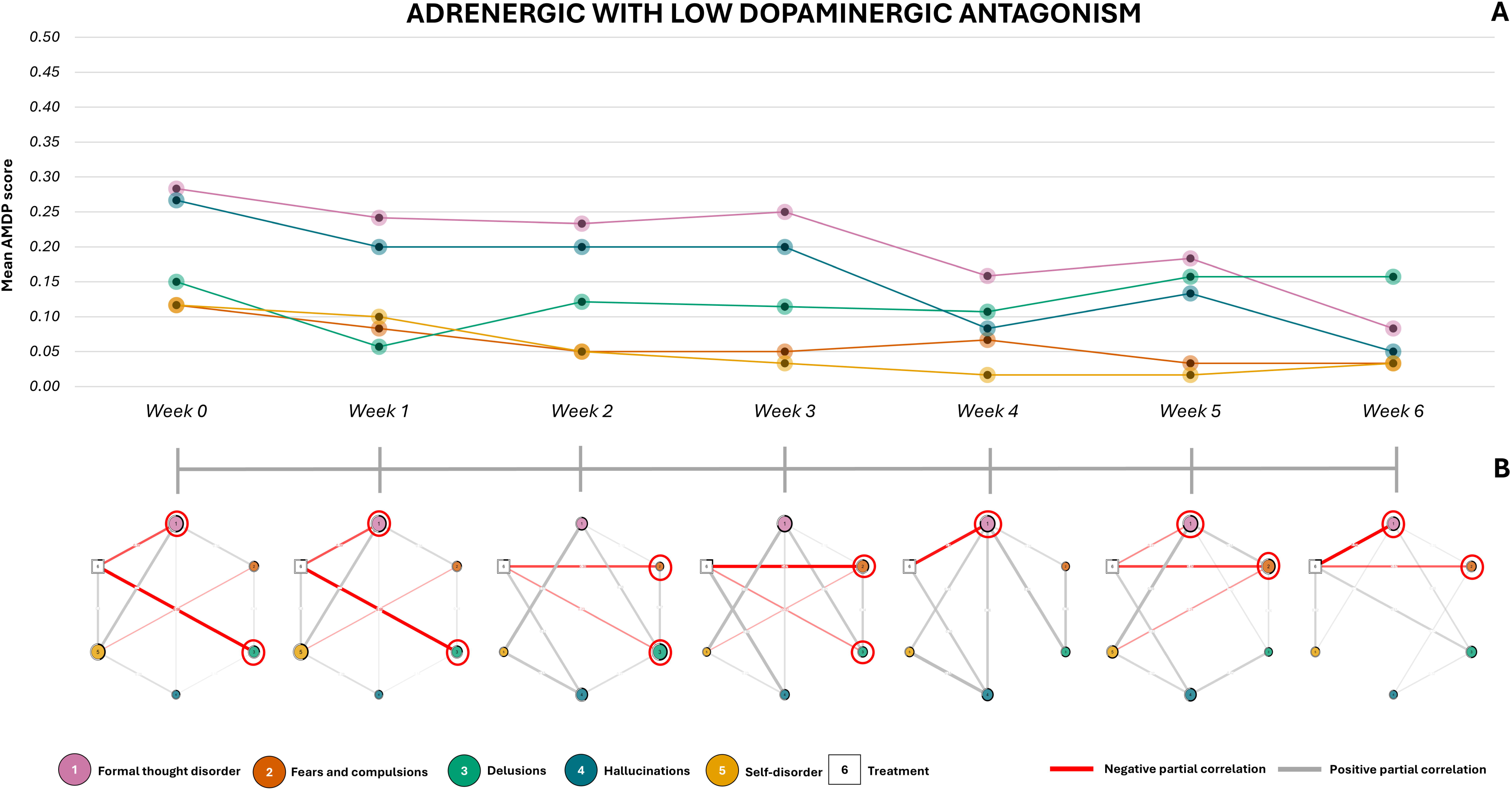
Part A, line graph showing the development of symptoms over time for the Manual for the Assessment and Documentation of Psychopathology in Psychiatry (AMDP) from T0 to T6 (given as mean of the AMDP rating forms). Grey connections indicate positive partial correlations, while red connections indicate negative partial correlations. The ring around each node indicates its predictability, the darker the ring, the higher the value. The size of the nodes is proportional to their expected influence value. The colour of the nodes corresponds to that of the lines in the Part A graph. The white square node represents the pharmacological treatment. The red circles mark the symptom nodes upon which the pharmacological treatment exerts a direct effect, contributing to their reduction.

**Table 2:** The table presents the expected influence (EI) and predictability (PRD) for each symptom across the 7 time points. It also includes the network density index for each model within the three drug groups.

|  | Week 0 |  | Week 1 |  | Week 2 |  | Week 3 |  | Week 4 |  | Week 5 |  | Week 6 |  |
| --- | --- | --- | --- | --- | --- | --- | --- | --- | --- | --- | --- | --- | --- | --- |
|  | PRD | EI | PRD | EI | PRD | EI | PRD | EI | PRD | EI | PRD | EI | PRD | EI |
| Muscarinic |  |  |  |  |  |  |  |  |  |  |  |  |  |  |
| Formal thought disorder | 0.54 | 0.94 | 0.48 | 0.96 | 0.60 | 1.00 | 0.71 | 1.00 | 0.81 | 1.00 | 0.71 | 0.92 | 0.51 | 1.00 |
| Fears and compulsions | 0.29 | 0.55 | 0.25 | 0.51 | 0.34 | 0.53 | 0.35 | 0.51 | 0.49 | 0.32 | 0.46 | 0.69 | 0.42 | 0.68 |
| Delusions | 0.29 | 0.79 | 0.23 | 0.66 | 0.51 | 0.78 | 0.57 | 0.78 | 0.67 | 0.61 | 0.52 | 0.84 | 0.51 | 0.93 |
| Hallucinations | 0.40 | 0.75 | 0.27 | 0.56 | 0.39 | 0.70 | 0.40 | 0.36 | 0.70 | 0.42 | 0.52 | 0.57 | 0.33 | 0.50 |
| Self-disorder | 0.49 | 1.00 | 0.44 | 1.00 | 0.56 | 0.91 | 0.55 | 0.65 | 0.61 | 0.42 | 0.64 | 1.00 | 0.12 | 0.16 |
| Mean | 0.40 | \ | 0.33 | \ | 0.48 | \ | 0.51 | \ | 0.65 | \ | 0.57 | \ | 0.38 | \ |
| Network Density | 0.80 |  | 0.87 |  | 0.67 |  | 0.60 |  | 0.53 |  | 0.60 |  | 0.53 |  |
| Dopaminergic/Serotonergic |  |  |  |  |  |  |  |  |  |  |  |  |  |  |
| Formal thought disorder | 0.47 | 1.00 | 0.47 | 1.00 | 0.59 | 0.86 | 0.71 | 1.00 | 0.79 | 1.00 | 0.71 | 1.00 | 0.49 | 1.00 |
| Fears and compulsions | 0.25 | 0.73 | 0.25 | 0.73 | 0.36 | 0.44 | 0.33 | 0.51 | 0.49 | 0.38 | 0.46 | 0.75 | 0.40 | 0.55 |
| Delusions | 0.21 | 0.65 | 0.21 | 0.65 | 0.52 | 0.96 | 0.58 | 0.87 | 0.66 | 0.81 | 0.52 | 0.80 | 0.47 | 0.73 |
| Hallucinations | 0.27 | 0.70 | 0.27 | 0.70 | 0.44 | 1.00 | 0.43 | 0.69 | 0.70 | 0.75 | 0.54 | 0.80 | 0.33 | 0.69 |
| Self-disorder | 0.41 | 0.93 | 0.41 | 0.93 | 0.55 | 0.92 | 0.59 | 0.66 | 0.60 | 0.50 | 0.63 | 0.82 | 0.15 | 0.55 |
| **Mean** | 0.32 | \ | 0.32 | \ | 0.49 | \ | 0.53 | \ | 0.65 | \ | 0.57 | \ | 0.37 | \ |
| **Network Density** | **0.73** |  | **0.73** |  | **0.60** |  | **0.70** |  | **0.66** |  | **0.73** |  | **0.60** |  |
| **Adrenergic/Low dopaminergic** |  |  |  |  |  |  |  |  |  |  |  |  |  |  |
| Formal thought disorder | 0.50 | 1.00 | 0.50 | 1.00 | 0.59 | 0.87 | 0.71 | 1.00 | 0.79 | 1.00 | 0.71 | 1.00 | 0.56 | 0.71 |
| Fears and compulsions | 0.24 | 0.54 | 0.21 | 0.54 | 0.34 | 0.75 | 0.36 | 0.86 | 0.49 | 0.34 | 0.47 | 0.91 | 0.41 | 0.42 |
| Delusions | 0.25 | 0.68 | 0.25 | 0.68 | 0.51 | 1.00 | 0.55 | 0.74 | 0.64 | 0.44 | 0.54 | 0.59 | 0.50 | 0.50 |
| Hallucinations | 0.25 | 0.42 | 0.25 | 0.42 | 0.40 | 0.89 | 0.44 | 0.73 | 0.71 | 0.65 | 0.55 | 0.81 | 0.33 | 0.22 |
| Self-disorder | 0.45 | 0.97 | 0.45 | 0.97 | 0.55 | 0.77 | 0.54 | 0.66 | 0.65 | 0.44 | 0.63 | 0.79 | 0.18 | 0.33 |
| **Mean** | 0.34 | \ | 0.33 | \ | 0.48 | \ | 0.52 | \ | 0.66 | \ | 0.58 | \ | 0.40 | \ |
| **Network Density** | **0.66** |  | **0.66** |  | **0.60** |  | **0.66** |  | **0.53** |  | **0.66** |  | **0.60** |  |

### 3.1 Patients undergoing treatment with muscarinic M2-M5 antagonists

The temporal network analysis presented in Fig. 1B provides insight into the psychopathological changes that occurred by identifying specific muscarinic treatment targets at different time points. During the first two weeks, treatment primarily targeted self-disorder, progressively reducing its influence within the network (EI_T0_= 1; EI_T6_= 0.14). Given the positive associations between self-disorder and both formal thought disorder and hallucinations, this attenuation contributed to the overall and consistent symptom reduction observed in the line graphs.

From T3 onwards, the primary treatment target shifted from self-disorder to delusions, which had gained prominence within the network due to their increasing expected influence (EI_T0_ = 0.29; EI_T3_ = 0.57). The decrease observed for both formal thought disorder, and hallucinations is the result of an indirect action of the drug mediated firstly by self-disorder and then by delusions. As shown in Table 2, from T2 onwards, the network displayed a less interconnected structure with a density index of 0.53 at T6, suggesting a potential reduction in symptom co-dependence over time and the transition to a more resilient state. Furthermore, the mean predictability values (i.e., the amount of variance explained by other symptoms and treatment) for each variable increase between T2 and T4, reaching an average of up to 65% of the symptom variance.

### 3.2 Patients undergoing treatment with serotoninergic and dopaminergic antagonists

Network analysis (Fig. 2B) revealed that the treatment with serotoninergic and dopamine antagonists consistently targeted hallucinations across time points. At T1 and T2 it also directly affected fears and compulsions, while at T4, the primary target shifted to formal thought disorder. By T6, the treatment showed a strong connection with self-disorder, which remained positively linked to formal thought disorder.

As for the previous group, network nodes’ predictability increased between T0 and T4, reaching 65% of explained variance. This was accompanied by a reduction in connection density, resulting in networks that were more resilient to change. Notably, at T5, when all symptoms worsened compared to the previous time point, the network showed higher connection density (0.73), and treatment had the fewest and weakest negative connections. At T6, the network density decreased again (0.60), the treatment has two direct connections with hallucinations and self-disorder and as can be seen from Fig. 2A, the symptoms had again decreased compared to T5.

The most influential node in the network across all time points was always formal thought disorder (EI = 1). T5 was the only time point at which the treatment targeted this symptom. The node with the second highest EI value at T0 (self-disorder) showed a decrease in its EI throughout the treatment (EI_T0_ = 0.93, EI_T0_ = 0.55).

### 3.3 Patients undergoing treatment with adrenergic agents with low dopaminergic antagonism

The network models obtained for the group treated with adrenergic agents with low dopaminergic antagonism and depicted in Figure 3B, show that the treatment node primarily targeted formal thought disorder and delusions during the first two time points. At T2 and T3 the connections shifted to fears and compulsions and delusions and then target formal thought disorder and fears and compulsions for the remaining period.

Symptom predictability peaked at T4, explaining 66% of the variance in the network nodes. However, connection density indices remained stable and were comparable to the final values observed in the other two groups. Additionally, formal thought disorder, the symptom with the highest EI values at T0, was targeted in five of seven time points. Delusions and fears and compulsions became targets of the treatment node as they gained greater importance within the network. The EI values for each time point and node are reported in Table 2.

## 4. Discussion

The aim of this study was to examine how different antipsychotic treatments influence the evolving interplay between psychotic symptoms over time, by applying NIA to model treatment as an active node within the network. By tracking changes across seven weekly time points and classifying antipsychotics based on their receptor affinity profiles, we aimed to capture both cross-sectional and longitudinal variations in treatment response.

Our findings revealed distinct, time-dependent treatment targets for each drug class. Muscarinic antagonists initially targeted self-disorder, but as treatment progressed, the focus shifted to delusions. In contrast, serotonergic-dopaminergic antagonists primarily influenced hallucinations, with a notable rebound in symptoms at later stages. Adrenergic agents with low dopaminergic antagonism demonstrated a stabilising effect. These findings move beyond static measures of efficacy, providing a dynamic, network-level perspective on antipsychotic action to highlight how treatments interact with the broader architecture of symptom relations. Rather than examining symptoms in isolation, this approach emphasises how certain nodes may exert disproportionate influence within the symptom network, sustaining or amplifying others. Recognising these dynamics across treatment trajectories opens the way for more targeted and potentially synergistic interventions, tailored not only to the pharmacological profile of the drug but also to the evolving symptom interrelations in each patient.

### 4.1 Pharmacological profiles and network-based mechanisms of symptom change

Across the three pharmacological profiles studied, muscarinic antagonists emerged as the most effective in producing sustained symptom reduction and subsequent network stabilisation. This is evidenced by the consistently decreasing symptom trajectories compared to the other two drug classes and network dynamics over time. The networks became progressively less dense and more predictable, suggesting a coherent and sustained therapeutic response with increased system resilience (Cramer et al., 2016). These findings suggest that muscarinic antagonists may exert therapeutic effects not only by reducing symptom severity but also by weakening maladaptive symptom interconnections. This aligns with network-based models of psychopathology, which propose that tightly connected symptom networks can sustain chronicity and increase the risk of relapse (Scheffer et al., 2024).

Broader meta-analytic evidence supports our finding of differential efficacy of various antipsychotic classes. Leucht et al. (2023) showed that muscarinic antagonists like olanzapine rank among the most effective for reducing overall psychotic symptoms across different treatment durations. Moreover, longitudinal studies indicate that the majority of symptomatic improvement tends to occur within the first 4 - 6 weeks of treatment—often referred to as the “early phase”—after which further gains typically plateau (Keepers et al., 2020).

Serotoninergic-dopaminergic antagonists were associated with an almost exclusive focus on hallucinations, but, overall, with a less clearly defined decrease in symptom severity. This indicates a general but less clearly defined symptom reduction over the six weeks, especially when compared to the more consistent trajectory observed with muscarinic antagonists. Moreover, the apparent increase in all AMDP mean scores at T5 corresponds with a phase of network destabilisation, marked by a higher density of connections. Since all symptoms are positively related—meaning an increase in one tends to amplify the others—this rise in connectivity likely contributed to heightened overall symptom activation. This increased symptom interdependence may reflect a failure to sufficiently disentangle individual symptom dynamics during this phase.

Adrenergic agents with low dopaminergic antagonism showed minimal symptom reduction but uniquely maintained stable effects across all seven time points. They consistently targeting the most influential nodes in the networks, as reflected by the relatively flat symptom trajectory in Figure 3A. This suggests that such treatments are more associated with sustaining symptom stability than promoting symptom improvement.

The evidence of drug class-specific symptom targets becomes even more compelling when considered alongside the pharmacological profile of clozapine. Although not included in this study, clozapine’s unique receptor binding—spanning muscarinic, serotonergic, and dopaminergic systems—has long been associated with its superior efficacy, particularly in treatment-resistant schizophrenia (Correll, 2025). This profile offers a valuable benchmark for contextualising our findings, highlighting how targeting multiple pathways may produce broader clinical effects. This consideration supports a model where multi-target pharmacodynamics enable more extensive network reconfiguration and symptom disentanglement.

Our observations resonate with previous network analyses’ findings that emphasise the importance of studying symptom dynamics. For instance, our results align with those of Sun et al. (2023), who reported a progressive reduction in the strength and density of symptom interconnections over the course of treatment. This decrease was associated with greater network resilience, indicating that effective pharmacological interventions may work not only by reducing symptom severity but also by decoupling tightly knit symptom clusters. Conversely, Zamani et al. (2017) reported an increase in network density and symptom centrality following treatment. Methodological differences may account for this inconsistency. Their analysis was based on two time points—pre- and post-treatment—offering a more static snapshot of network change. In contrast, our longitudinal approach reveals that network dynamics evolve in non-linear ways across multiple phases of treatment. For example, while symptom predictability increased steadily over the first four weeks (explaining up to 65–66% of variance), reductions in network density were more gradual and treatment-specific.

### 4.2 Clinical implications

This study’s findings highlight that effective treatment may depend not only on reducing symptom severity but also on modifying how symptoms influence one another over time. In this framework, therapeutic success is tied to decoupling maladaptive symptom patterns and promoting more adaptive network configurations. Monitoring the centrality and connectivity of symptoms can provide clinicians with valuable insights, helping to identify what to prioritise and at what time. Analysing the evolving structure of the symptom network may also guide the more tailored integration of pharmacological and psychotherapeutic treatments. For example, during early treatment phases, medications may successfully suppress core symptoms such as self-disorder or hallucinations. However, other nodes—such as formal thought disorder or emerging delusional content—may remain less affected or gain importance. In such cases, psychological interventions could be strategically employed to target these residual or ascending domains, complementing the pharmacological action.

Beyond enhancing therapeutic targeting, time-resolved network models may also provide early warnings of adverse effects linked to specific pharmacodynamic profiles. Positive associations between treatment and certain symptoms—manifesting as grey edges in the NIA—may signal emerging side effects or compensatory mechanisms. For example, in Figure 1B, muscarinic antagonists positive correlated with formal thought disorder and fears and compulsions. This pattern may reflect known anticholinergic-related cognitive impairments and anxiogenic effects, possibly mediated through indirect modulation of serotonergic and dopaminergic pathways (Guo et al., 2010; Haram et al., 2019). Recognising such vulnerabilities may facilitate anticipatory adjustments in treatment.

### 4.3 Limitations

This study has some limitations that should be acknowledged. First, baseline data were collected after an initial stabilisation period of at least two weeks. This was necessary to ensure the feasibility and reliability of neuropsychological and clinical assessments. However, it limits the ability to capture the earliest effects of antipsychotic interventions on symptom networks. The observed dynamics therefore reflect the later phase of clinical improvement, when acute symptoms have largely remitted, and more subtle treatment effects emerge.

Second, antipsychotic treatment allocation was not randomised. Medications were prescribed based on clinical judgement in the context of routine care, potentially introducing selection biases related to individual clinical characteristics. This limits the ability to infer causal relationships between specific treatments and symptom changes. However, the use of NIA in this context highlights its potential applicability to other treatment phases, including acute settings, and supports its future integration into randomised controlled trials to more robustly evaluate class-specific treatment effects.

Lastly, the pharmacological groups were not balanced in terms of sample size. Although this reflects the naturalistic setting of the study, it may have led to unequal statistical power across comparisons and could limit the generalisability of some findings.

### 4.4 Conclusions

Our findings revealed differential patterns of symptom network evolution under distinct pharmacological profiles. By identifying dynamic treatment targets, we highlight the value of examining symptom trajectories within a network framework rather than focusing solely on total symptom severity.

The evolving architecture of symptom interactions provides clinically relevant insights that extend beyond efficacy. Network modelling can help anticipate vulnerabilities and emerging side effects linked to specific receptor profiles, contributing to more informed and adaptive treatment strategies. These results support the development of phase-specific, network-informed interventions that combine pharmacological and psychological approaches in a more integrated and personalised manner. Future research should replicate and expand on these findings in larger, more diverse samples, with earlier assessment time points to fully capture the initial impact of treatment on symptom dynamics.

## Data Availability

All data produced in the present study are available upon reasonable request to the authors

## Author Disclosures

### Financial support

The STRICON project was supported by a grant from the Brain & Behavior Research Foundation (Grant No. 28445), awarded to Philipp Homan, MD, PhD.

### Author Contributions

Conceptualisation, P.S., G.C., J.M.C.B., and P.H.; methodology, P.S., G.C., and J.M.C.B.; validation, P.S., and G.C.; formal analysis, P.S., and J.M.C.B.; Investigation, G.C.; resources, P.H.; data curation, P.S., G.C., W.O.; writing—original draft preparation, P.S., G.C, V.E.; writing—review and editing, P.S., G.C., V.E., and J.M.C.B.; supervision, P.H., W.O.; project administration, G.C. All authors have read and agreed to the published version of the manuscript.

### Declaration of Interest

The authors declare no competing interests

### Ethical statement

The authors assert that all procedures contributing to this work comply with the ethical standards of the relevant national and institutional committees on human experimentation and with the Helsinki Declaration of 1975, as revised in 2013. All procedures involving human subjects/patients were approved by the local ethics committee (KEK-ZH 2020/01049)

## Acknowledgements

The authors would like to thank the Società Italiana di Farmacologia (SIF) for supporting this research by awarding the first author, P. Sarti with the SIF Research Grant for Short Periods Abroad 2024-2025

## Notes

### Competing Interest Statement

The authors have declared no competing interest.

### Author Declarations

The authors assert that all procedures contributing to this work comply with the ethical standards of the relevant national and institutional committees on human experimentation and with the Helsinki Declaration of 1975, as revised in 2013. All procedures involving human subjects/patients were approved by the local ethics committee (Ethics Committee/IRB of the Kantonale Ethikkommission Zürich (Stampfenbachstrasse 121, 8090 Zürich) - Project ID: KEK-ZH 2020-00277)

